# Enhancing immunotherapy outcomes in glioblastoma through predictive machine learning

**DOI:** 10.1101/2023.10.05.23296617

**Authors:** Guillaume Mestrallet

## Abstract

Glioblastoma is a highly aggressive cancer associated with a dismal prognosis, with a mere 5% of patients surviving beyond five years post-diagnosis. Current therapeutic modalities encompass surgical intervention, radiotherapy, chemotherapy, and immune checkpoint inhibitors (ICB). However, the efficacy of ICB remains limited in glioblastoma patients, necessitating a proactive approach to anticipate treatment response and resistance.

In this comprehensive study, we conducted a rigorous analysis involving two distinct glioblastoma patient cohorts subjected to PD-1 blockade treatments. Our investigation unveiled that a significant portion, 60%, of patients exhibit persistent disease progression despite ICB intervention. To elucidate the underpinnings of resistance, we characterized the immune profiles of glioblastoma patients with continued cancer progression following anti-PD1 therapy. These profiles revealed multifaceted defects, encompassing compromised macrophage, monocyte, and T follicular helper responses, impaired antigen presentation, aberrant regulatory T cell (Tregs) responses, and heightened expression of immunosuppressive molecules (TGFB, IL2RA, and CD276). Building upon these resistance profiles, we leveraged cutting-edge machine learning algorithms to develop predictive models and accompanying software. This innovative computational tool achieved remarkable success, accurately forecasting the progression status of 82.82% of glioblastoma patients following ICB, based on their unique immune characteristics.

In conclusion, our pioneering approach advocates for the personalization of immunotherapy in glioblastoma patients. By harnessing patient-specific attributes and computational predictions, we offer a promising avenue for the enhancement of clinical outcomes in the realm of immunotherapy. This paradigm shift towards tailored therapies underscores the potential to revolutionize the management of glioblastoma, opening new horizons for improved patient care.

## Introduction

Glioblastoma is the most common primary malignant central nervous system tumor, with an incidence rate of 3.19 per 100,000 persons in the United States of America (1,2). The median survival is only 15 months, the median progression-free survival is 7 months, and less than 5% of the patients survive 5 years following diagnosis.

Therapeutic options include surgery, chemotherapy (temozolomide), radiotherapy and immune checkpoint blockade (ICB) (2,3). However, most of the patients remain resistant to PD-1 blockade in the adjuvant settings, except for patients with mismatch repair deficiency (2,4–10). Importantly,pPatients who received neoadjuvant pembrolizumab with continued adjuvant therapy following surgery had significantly extended overall survival compared to patients that received adjuvant, post-surgical anti-PD-1 alone (2). It was associated with upregulation of T cell and IFNγ related gene expression and downregulation of cell-cycle related gene expression within the tumor, fewer monocytes in the blood compared with patients that received adjuvant therapy. However, even in neoadjuvant settings, patient resistance to ICB still occurs.

Thus, conducting a meta-analysis of glioblastoma patient cohorts will be instrumental in characterizing the mechanisms underpinning response and resistance to ICB. Meta-analysis of data from multiple cohorts may also facilitate the identification of optimal targets to develop combination therapies and improve patient outcomes (2,5). The development of software using machine learning approaches will enhance the precision of response and resistance prediction to ICB. It will improve the diagnosis and subsequent therapeutic strategies according to patient-specific characteristics.

## Material and methods

### RNAseq datasets and selection of cohorts

Patient cohort was selected using the CRI iAtlas Portal (11). We selected the following RNAseq datasets for glioblastoma patients : Zhao 2019 - GBM, PD-1 (5), Prins 2019 - GBM, PD-1 (2). We used the following group filters : Progression, Drug and GBM. Non-Progressors are defined as patients with mRECIST of Partial Response, Complete Response or Stable disease, whereas Progressors are those with Progressive Disease. Then, we used the ICI Analysis Modules. The current version of the iAtlas Portal was built in R using code hosted at https://github.com/CRI-iAtlas/iatlas-app. Assayed samples were collected prior to immunotherapy.

### Clinical description of patients

Merged datasets according to drug therapy are described in **Table 1**. For patients that received Nivolumab, targeting PD1, there are 11 (40%) non progressors and 17 (60%) progressors. For patients that received Pembrolizumab, targeting PD1, there are 15 (44%) non progressors and 19 (56%) progressors.

**Table 1).** Response of patients with glioblastoma to PD1 blockade.

| Drug | Nivolumab | Pembrolizumab |
| --- | --- | --- |
| Target | PD1 | PD1 |
| Non progressors (nb) | 11 | 15 |
| Progressors (nb) | 17 | 19 |
| Non progressors (%) | 40 | 44 |
| Progressors (%) | 60 | 56 |

### Immune landscape of cancer in iAtlas

The initial release of iAtlas provided a resource to complement analysis results from The Cancer Genome Atlas (TCGA) Research Network on the TCGA data set comprising over 10,000 tumor samples and 33 tumor types (The Immune Landscape of Cancer; here referred to as Immune Landscape) (12).

### Statistics

Statistical significance of the observed differences was determined using the independent Wilcoxon t-Test with multiple sample correction. All data are presented as mean±SEM. The difference was considered as significant when the p value was below 0.05. * : p<0.05.

### Software development to predict response to PD1 blockade

Patients from 2 cohorts received anti-PD-1 following glioblastoma. After pooling the cohorts, progression was calculated according to the drug used for therapy. n=28 for Nivolumab and n=34 for Pembrolizumab. The software, coded using python, html, css, mysql and django, allows the registered clinician to diagnose a new patient or access the diagnosis of a registered patient by indicating their medical identifier in a form, as we previously described in other studies (13,14). The software calculates the probability of patients to respond to anti-PD1 after form completion.

## Results

### Progression and overall survival of glioblastoma patients according to PD1 blockade in two cohorts

We calculated the progression and overall survival of glioblastoma patients who underwent immune checkpoint therapy targeting PD1 in two cohorts. We aggregated the results based on the progression status to each checkpoint combination (**Table 1**). For patients that received Nivolumab, targeting PD1, there are 11 (40%) non progressors and 17 (60%) progressors. For patients that received Pembrolizumab, targeting PD1, there are 15 (44%) non progressors and 19 (56%) progressors. For patients receiving monotherapy targeting PD1 with disease progression, overall survival remained below 30% (**Figure 1**). Conversely, non-progressors following Pembrolizumab exhibited an overall survival rate around 60%. Overall, 60% of patients with glioblastoma exhibited strong resistance to PD1 blockade therapy.

**Figure 1).**
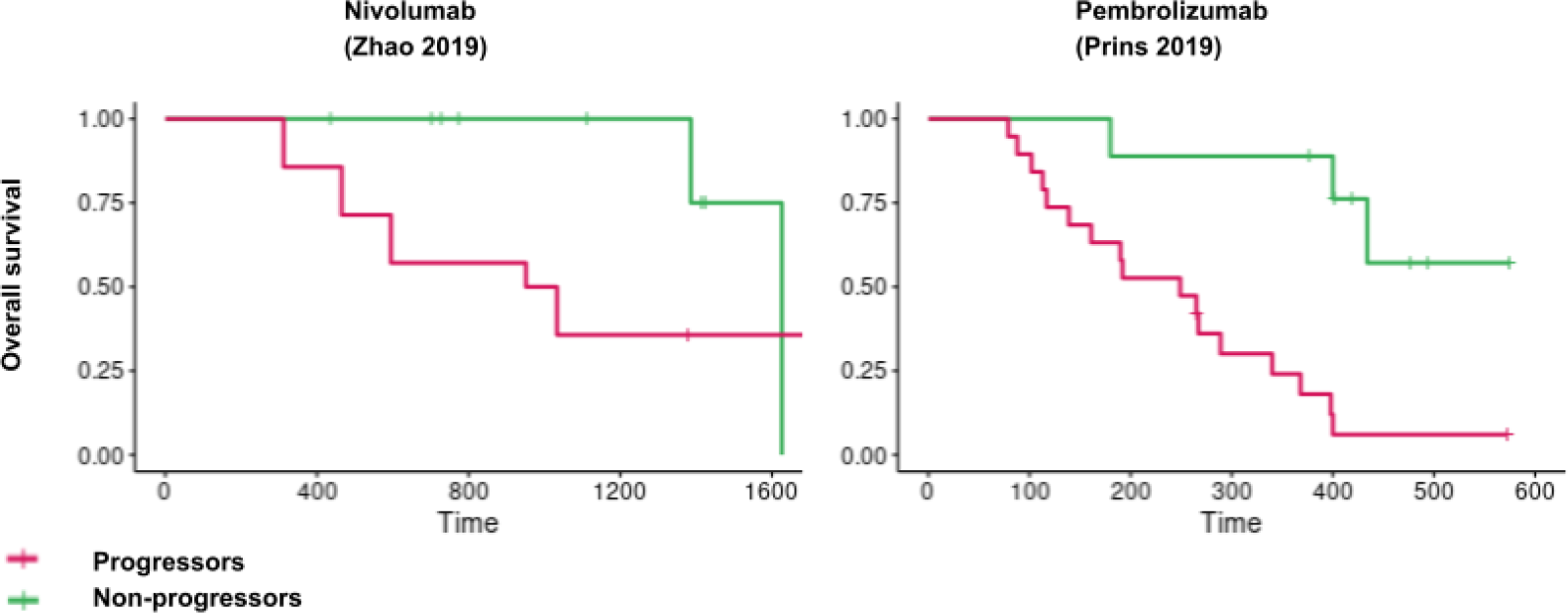
Overall survival of glioblastoma patients according to PD1 blockade in 2 cohorts. Patients from 2 cohorts received anti-PD-1 following glioblastoma. Overall survival was calculated in these cohorts. n=28 for Nivolumab and n=34 for Pembrolizumab.

### Immune response and resistance in glioblastoma patients following PD1 blockade

We investigated the immune features associated with response and resistance to immune checkpoint therapy in glioblastoma patients. Analyzing immune response using the CRI iAtlas in patients who received anti-PD1 (Nivolumab or Pembrolizumab), we observed that non-progressors to Pembrolizumab had more monocytes and T follicular helpers, while progressors had more macrophages, especially M0, and Tregs (**Figure 2**). No significant differences were observed for other immune subsets in the Pembrolizumab cohort, and no difference at all in the Nivolumab cohort (**Figure 2, Supplementary figure 1**).

**Figure 2).**
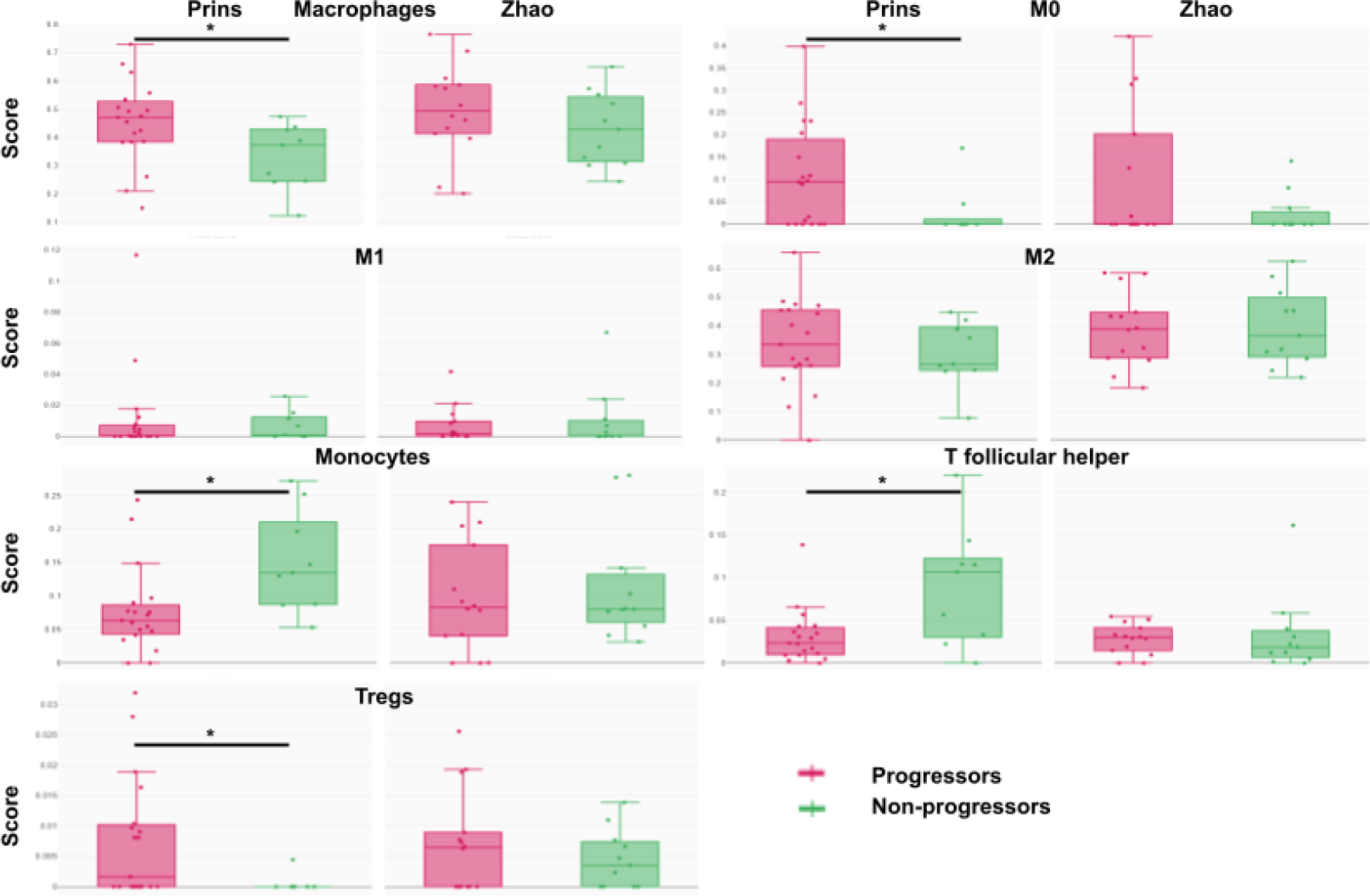
Immune response in glioblastoma patients according to progression following PD1 blockade. 62 patients received anti-PD-1 following glioblastoma. n=28 for Nivolumab (Zhao) and n=34 for Pembrolizumab (Prins). Immune response is measured using CRI iAtlas. p<0.05, Wilcoxon t-test.

When examining the expression of immunomodulatory molecules, we found that progressors following Pembrolizumab expressed more MHC molecules (HLA-DRB1, HLA-DQA1, HLA-DRB5, HLA-DQB1) compared to non-progressors. This suggests that defects in antigen presentation plays a role in the anti-tumor immune resistance (**Figure 3**). Surprisingly, we observed upregulation of ITG2B, a gene related to T cell adhesion in progressors following Pembrolizumab, suggesting that T cell responses are not sufficient to promote response. Importantly, progressors following Pembrolizumab express more immunosuppressive molecules such as TGFB, IL2RA and CD276. Progressors following Nivolumab exhibited lower expression of BTN3A1, a gene involved in T cell activation, IL4 and ARG1. No differences were observed in the expression of other immunoregulatory molecules and immune checkpoints (including PD1, TIGIT, TIM3, LAG3, EDNRB, TLR4, VSIR, CD40, TNFRSF, CD28, ICOS, VTCN1, CD70, CX3CL1, ENTPD1, GXMA, HMGB1, ICOSLG, VEGF, KIR, IFN genes, interleukins, MICA, and other HLA genes) (**Supplementary figure 2**). Of note, the differences observed in one cohort were not observed in the other cohort (**Figure 1/2, Supplementary Figures 1/2**). Overall, glioblastoma patients with cancer progression following anti-PD1 therapy were characterized by defects in macrophage, monocyte and T follicular helper responses, impaired antigen presentation, Tregs response and immunosuppressive molecule expression (TGFB, IL2RA and CD276).

**Figure 3).**
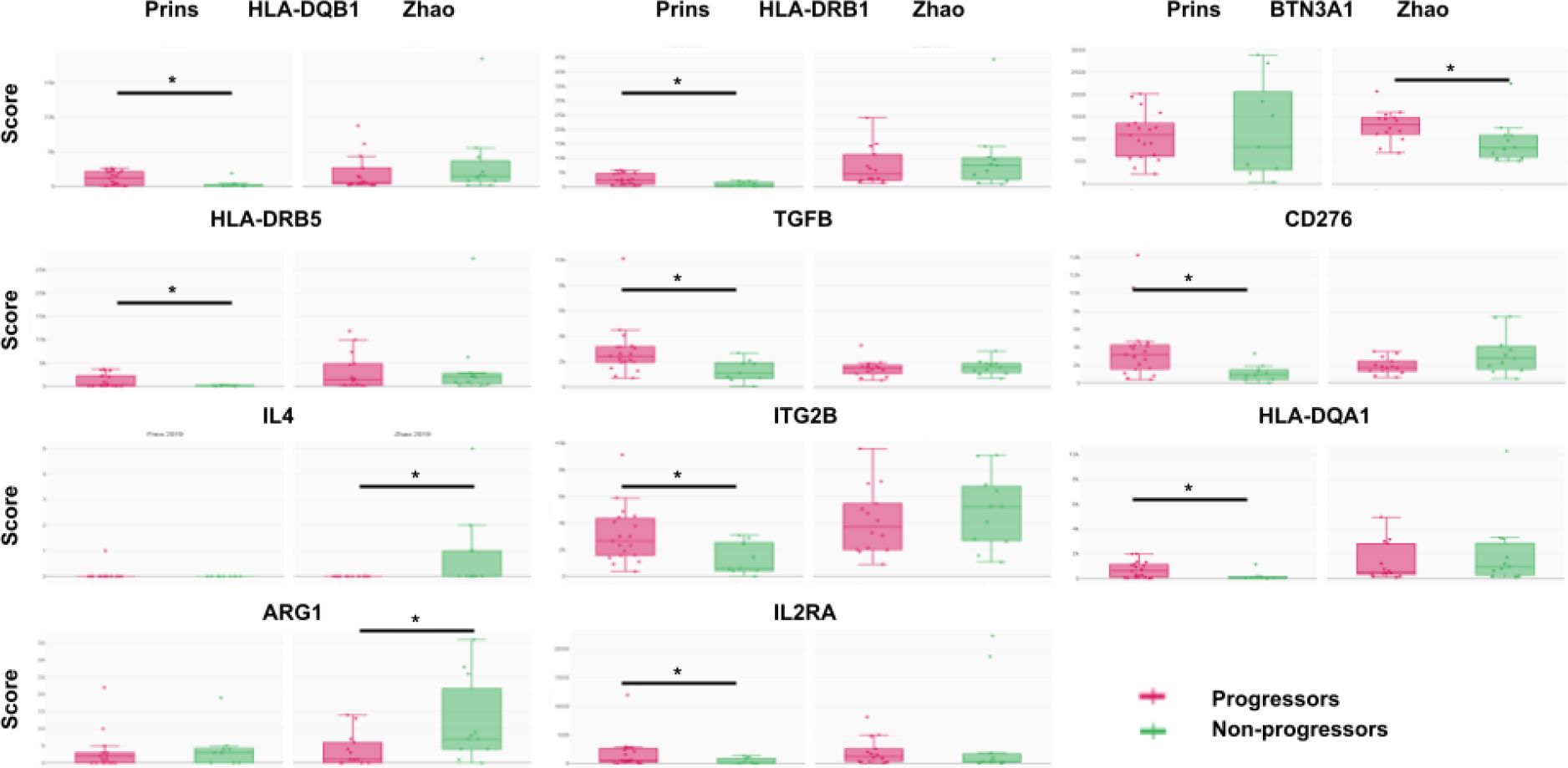
Immunomodulatory molecule expression in glioblastoma patients according to progression following PD1 blockade. 62 patients received anti-PD-1 following glioblastoma. n=28 for Nivolumab (Zhao) and n=34 for Pembrolizumab (Prins). Immunomodulatory molecule expression is measured using CRI iAtlas. p<0.05, Wilcoxon t-test.

### Software prediction of glioblastoma patient response to PD1 blockade

To predict the personalized response of patients with glioblastoma to PD1 blockade, we trained a RandomForestClassifier on the dataset, based on the features identified that are differentially expressed between Progressors and Non-progressors after PD1 blockade (**Figures 2/3**). The overall accuracy of our model is approximately 81.82%, which means that it correctly predicted the Progression status for about 81.82% of the data points in the test set (**Figure 4**). The classification report provides more detailed performance metrics, including precision, recall, and F1-score for each class (‘Non_Progressor’ and ‘Progressor’). Precision measures the accuracy of positive predictions. For ‘Non_Progressor,’ the precision is 100%, indicating that all positive predictions for this class were correct. For ‘Progressor,’ the precision is 80%, meaning that 80% of the positive predictions for this class were accurate. Recall measures the ability of the model to identify all relevant instances of a class. For ‘Non_Progressor,’ the recall is 33%, indicating that only 33% of the actual ‘Non_Progressor’ instances were correctly identified. For ‘Progressor,’ the recall is 100%, meaning that all actual ‘Progressor’ instances were correctly identified. The F1-score is the harmonic mean of precision and recall and provides a balance between the two. For ‘Non_Progressor,’ the F1-score is 0.50, and for ‘Progressor,’ it is 0.89. Support represents the number of samples in each class in the test set. For ‘Non_Progressor,’ there are 3 samples, and for ‘Progressor,’ there are 8 samples. Overall, our model managed to successfully predict the Progression status of 82,82% of the patients.

In accordance with the methods outlined in the study, we conducted predictions regarding the response of a hypothetical patient to PD1 blockade (**Figure 5**). Based on her genetic characteristics, the algorithm predicts that the patient has a 82,82% probability to be a Progressor following anti-PD1 therapy. Thus, it may be better to choose a different therapy for this patient.

**Figure 4).**
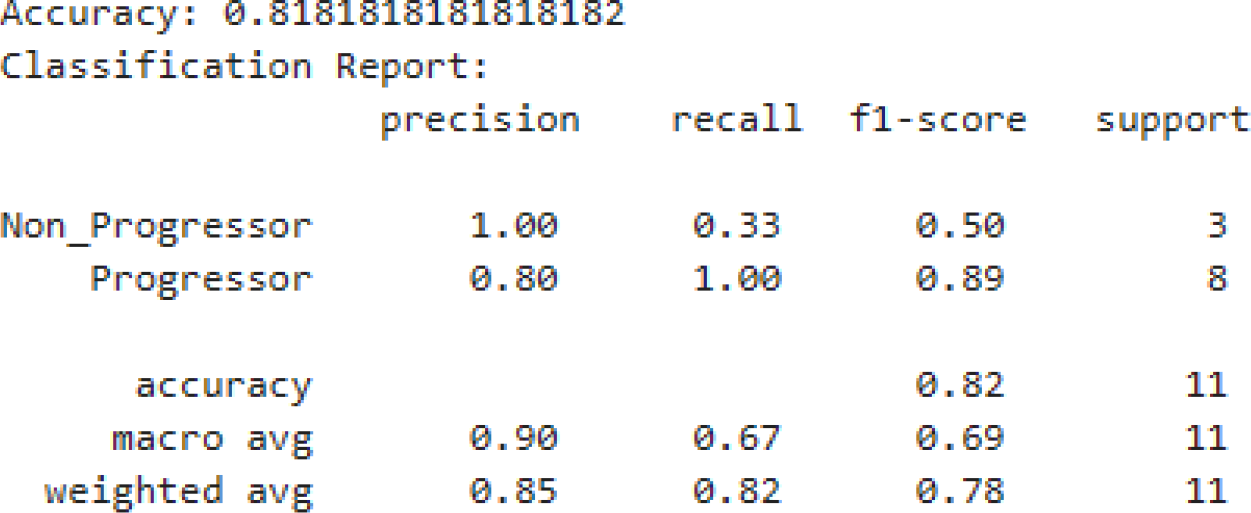
Performance of the algorithm predicting glioblastoma patient response to PD1 blockade.

**Figure 5).**
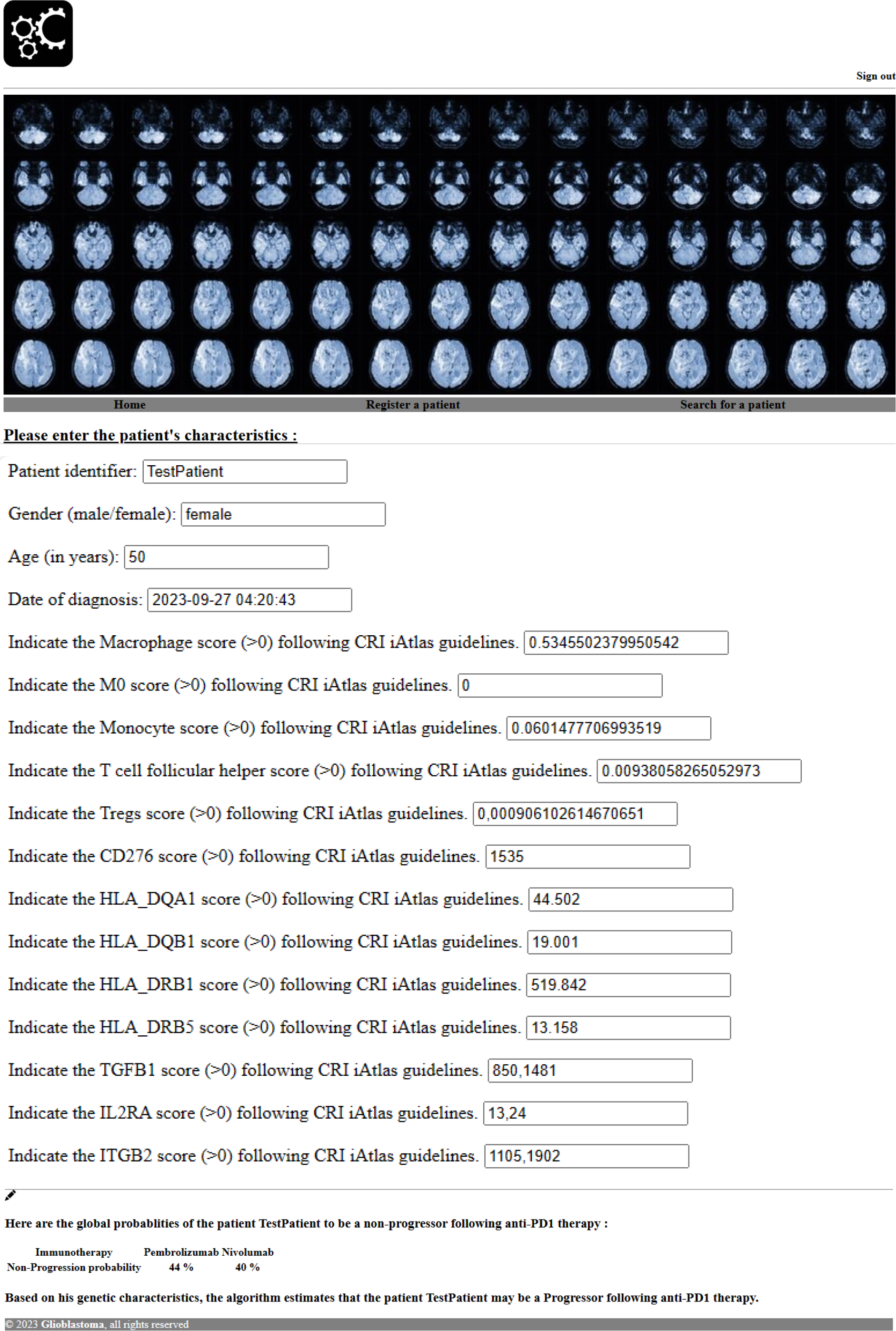
Software prediction of glioblastoma patient response to PD1 blockade. Glioblastoma patient data collection form. Global probability of the patient to respond to different immune checkpoint blockades. Personalized probability of the patient to respond to immune checkpoint blockade based on her genetic characteristics.

## Discussion

By analyzing the progression of patients following ICB in 2 cohorts, we observed that 60% of patients with glioblastoma exhibited strong resistance to PD1 blockade therapy. Glioblastoma patients with cancer progression following anti-PD1 therapy were characterized by defects in macrophage, monocyte and T follicular helper responses, impaired antigen presentation, Tregs response and immunosuppressive molecule expression (TGFB, IL2RA and CD276). CD276 regulates cell proliferation, invasion, and migration in cancers (15). High expression of IL2RA, the alpha chain of the interleukin 2 receptor complex expressed on the surface of mature T cells, predicted worse survival outcomes in patients with pancreatic ductal adenocarcinoma (16). TGFB is a key mediator of many biological processes and was also associated with resistance to immunotherapy (17). Progressors following Nivolumab also exhibited lower expression of BTN3A1, a molecule that coordinates αβ and γδ T cells (18). Many clinical trials are still ongoing and may provide more precise insights regarding immune resistance following ICB in these patients (4).

To better predict the ICB response of patients with glioblastoma, we developed machine learning approaches. We successfully trained a RandomForestClassifier on the CRI iAtlas dataset, based on the features that we identified as differentially expressed between Progressors and Non-progressors after anti-PD-1 blockade. Our model managed to successfully predict the Progression status of 82,82% of the patients. This model appears to have good precision and recall for the ‘Progressor’ class, suggesting that it can effectively identify ‘Progressor’ cases. However, there is room for improvement in identifying ‘Non_Progressor’ cases, as indicated by the lower recall for that class. Increasing the size of the training dataset would help to improve the predictions of the model.

Finally, we developed a software to predict patient response to immune checkpoint blockade that incorporated our machine learning approach. This software computes the probability of being a Progressor or a Non-progressor to PD-1 blockade based on the patient’s specific immune characteristics. Developing such machine learning approaches based on patient characteristics may help provide more relevant treatment to each patient.

## Data Availability

All data are openly available on CRI iAtlas website.

https://isb-cgc.shinyapps.io/iatlas/

## Acknowledgements

The author thanks Dr A. Pierga for the logo of the software.

## Supplementary Figures

**Supplementary Figure 1).**
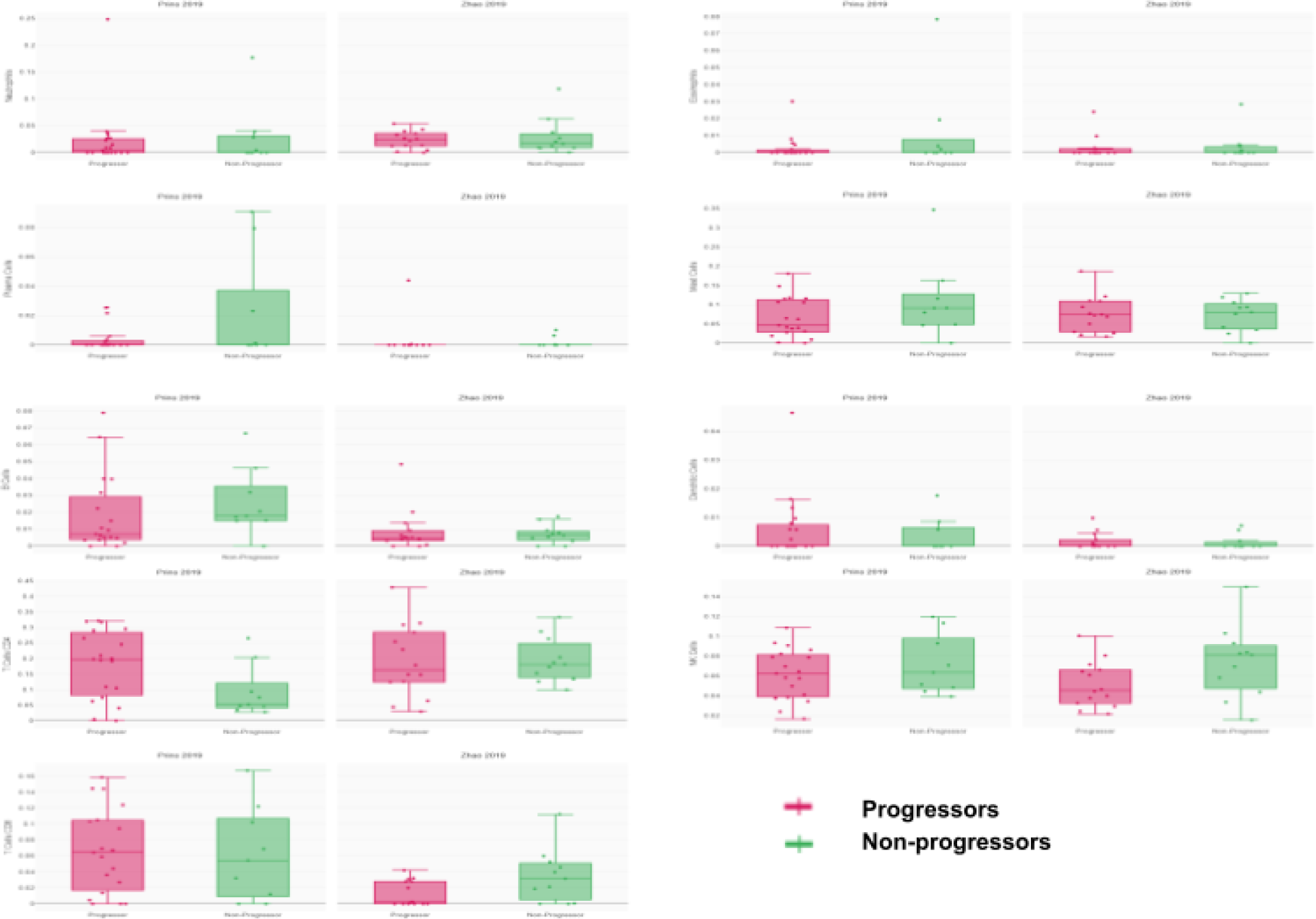
Immune response in glioblastoma patients according to progression following PD1 blockade. 62 patients received anti-PD-1 following glioblastoma. n=28 for Nivolumab (Zhao) and n=34 for Pembrolizumab (Prins). Immune response is measured using CRI iAtlas. p<0.05, Wilcoxon t-test.

**Supplementary Figure 2).**
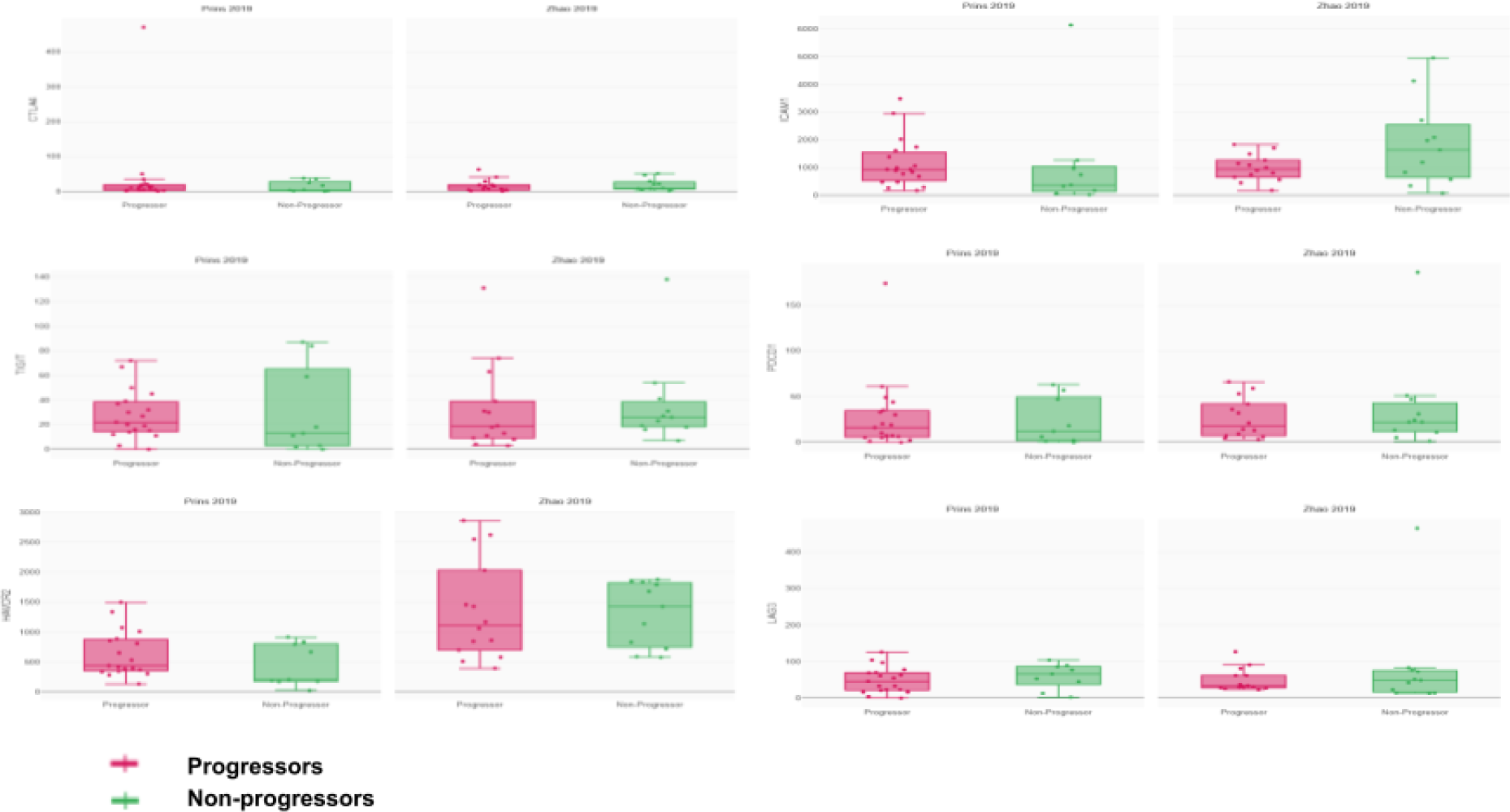
Immunomodulatory molecule expression in glioblastoma patients according to progression following PD1 blockade. 62 patients received anti-PD-1 following glioblastoma. n=28 for Nivolumab (Zhao) and n=34 for Pembrolizumab (Prins). Immunomodulatory molecule expression is measured using CRI iAtlas. p<0.05, Wilcoxon t-test.

